# The impact of a hybrid hospital at home program in reducing subacute rehabilitation referrals

**DOI:** 10.1101/2023.05.26.23290592

**Authors:** Ravi R Yadav, Mohammed A Mahyoub, Michael W Capriotti, Raul L Berio-Dorta, Kacie Dougherty, Ajit Shukla

**Affiliations:** Virtua Health, Marlton, NJ, USA; Systems Science and Industrial Engineering Department, Binghamton University, Binghamton, NY, USA

**Keywords:** managed care, NLP, text mining, short-term rehab, telemedicine

## Abstract

**Purpose:** The purpose of this study was to validate Hospital at Home as an appropriate care option for patients of certain diagnostic-related groups and acuity levels.

**Patients and methods:** We compared outcomes for patients in a Hospital at Home program at Virtua Health in 2022 (N = 272) to traditional inpatients at Virtua hospitals during the same year who did not participate in the Hospital at Home program (N = 13879). We defined outcomes as recommendations for subacute rehabilitation (SAR) and final disposition upon inpatient discharge. Specifically, we searched electronic medical records for terms related to recommendation for SAR and discharge to SAR using text mining algorithms and a natural language processing (NLP) model to confirm these recommendations.

**Results:** We observed that the proportion of patients in the Hospital at Home program that were recommended for SAR (0.147) was significantly different from the proportion of patients who remained in the hospital (0.361). Further, of those patients who received a recommendation, only 1 patient in the Hospital at Home group was discharged to SAR, while nearly half of those in the control group (proportion = 0.499) were transferred to SAR.

**Conclusion:** The Hospital at Home program is a promising alternative to traditional inpatient care for patients with certain diagnoses and who meet certain clinical criteria.

## Introduction

Healthcare digital transformation and telemedicine have noticeably increased during the past few years, especially with the COVID-19 pandemic^1–4^. Hospital at Home services, inpatient level of care in the home, began at Johns Hopkins University in the mid-1990s as a way to avoid the decline that many seniors experience as a component of care within hospital walls^5,6^. Throughout the years, several health systems adopted the model, but a broad payment structure did not exist making it financially challenging for more systems across the United States to fully implement similar programs^7^. In November 2020, the Centers for Medicare & Medicaid Services (CMS) launched the Acute Hospital Care at Home program to provide hospitals expanded flexibility to care for patients in their homes^8^. This unique waiver program was an expansion of the CMS Hospitals Without Walls legislation, as an effort to provide health systems with avenues to assist in treating the impending COVID-19 surges^9^. The result of this program was the first example of payment for this level of care at home in the United States^10^.

While the adoption of Hospital at Home programs across the United States had been slow prior to the CMS waiver program, the concept is not new internationally. For example, in Victoria, Australia upwards of six percent of all hospital bed-days are provided in the home^7^. In general, Hospital at Home programs have shown significant improvements in the quality of care including reduction in total length of stay, reduction in 30 day readmissions, reduced hospital infections and hospital induced delirium^11–13^(but see ^14,15^), as well as a lower total cost of care^13,16–19^(but see ^20^) and greatly improved patient satisfaction^11,15,16^.

Healthcare systems interested in starting virtual hospital programs are tasked with developing and deploying a model of care to meet patient needs, often involving frequent telehealth calls, at-home monitoring tools, and some in-home clinical care. Atrium Health, in the southeastern United States, described how they engaged multiple departments (hospital medicine, primary care, nursing, care management, research, administration, etc.) to develop a workflow that mimicked traditional patient care^21^. Patients in the program who qualified for traditional inpatient care received daily telehealth calls with a physician and twice daily telehealth calls from a nurse. Atrium Health also expanded community care to meet the higher demand for clinicians entering homes and delivered in-home monitoring kits to Hospital at Home patients.

Virtua Health applied for and was approved for the CMS waiver in September 2021 and launched its Hospital at Home program in January 2022 in response to a significant surge in hospital patients following the spread of the Omicron COVID-19 variant. Virtua’s Hospital at Home program began in one of its five hospitals and focused on transferring patients diagnosed with COVID-19 from a hospital floor to complete their care in a home bed. Today, Virtua’s Hospital at Home Program is available at all five hospitals with patients being transferred into the program from a hospital floor or directly from the emergency room after meeting admission criteria. The program includes more than 60 DRGs across several main DRG category codes including community acquired pneumonia, congestive obstructive pulmonary disease, congestive heart failure, COVID-19, cellulitis, and urinary tract infections. In 2022, Virtua’s Hospital at Home program treated more than 270 patients, with nearly 1,000 total inpatient days completed in a home bed.

Patients in Virtua’s Hospital at Home program receive support in the home from a well-integrated care team including physicians, nurses, and paramedics. Every Virtua Health Hospital at Home patient receives a remote patient monitoring kit which includes a tablet for virtual care visits via video as well as an armband continuously measuring heart rate, respiratory rate, body temperature, pulse oximetry, and step count. Patients also receive integrated peripheral devices depending on the diagnosis, such as a spirometer and weight scale. Patients are monitored and supported by Virtua’s 24/7 command center staffed by registered nurses. In addition, every patient receives two in-person nursing visits per day, a virtual physician visit from the attending hospitalists as well as six additional virtual rounding sessions with the command center registered nurses. Virtua’s program can support patients at home with virtual specialist consultations, oxygen concentrators, physical and occupational therapy, daily blood work and laboratory testing, mobile imaging including x-ray and ultrasound, oral and IV medications and fluids, as well as easy to heat nutritious meals.

The objective of this study was to validate the home as an appropriate care option for patients within certain DRGs and acuity levels. We wondered if care in the home would lead to overall improved health outcomes, possibly due to increased mobility and comfort in the home. We observed a significant difference in subacute rehabilitation (SAR) referrals between in-hospital and at-home patients, and suggest that this result supports future investigations into this alternative care model.

## Material and methods

### Study framework

In this study, we aimed to compare outcomes for patients who participated in Virtua’s Hospital at Home program to patients with the same DRGs who were not in the program. Specifically, we compared the number of patients recommended for SAR and the number of patients discharged to SAR in each group. Ethics approval was obtained from the Virtua Health IRB.

### Patient population

We limited the population in this study to patients who received inpatient-level hospital care at Virtua Health in 2022. To be admitted to the Hospital at Home program, patients needed to be on Medicare. We compared patients in Virtua’s Hospital at Home program (N = 272) to those patients who did not participate in the program (control, N = 13879). The control group was further limited to patients with a DRG that matched a DRG in the Hospital at Home patient group.

### Data collection

Patient data was recorded in an electronic medical record (EMR) using Epic (Epic, Epic Systems Corporation, Verona, Wisconsin, USA). To collect EMRs, we first queried Epic for patients in the Hospital at Home program using SQL. We then queried the database for EMRs for patients who did not participate in the program and with a DRG matching one of the DRGs in the Hospital at Home group. The following variables were extracted from the EMRs and saved in Excel files for use in further analysis: clinical note contents, discharge disposition, diagnosis related group, date of service, age (years) at admission, sex, and insurance payor. Information from clinical notes was extracted using natural language processing (NLP) and combined with tabular (structured) data for further analysis (**Figure 1**).

**Figure 1.**
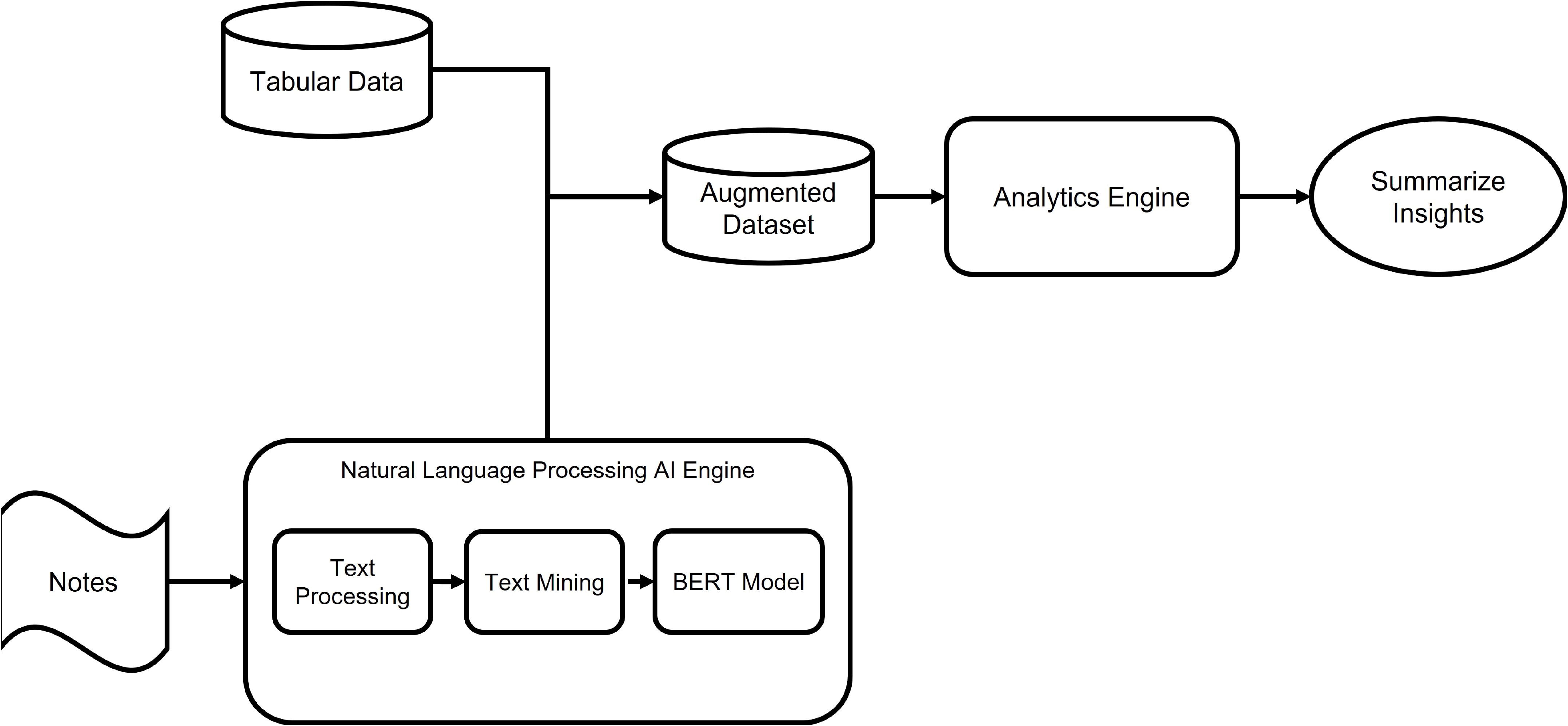
Flow diagram depicting the combination of clinical notes (unstructured data) and tabular (structured) data from EMRs used for analysis in this study. Clinical notes for each patient were collected from the EMR. The free text in the notes was processed (uniform capitalization, special characters removed) before each note was searched for keywords (see **Methods**). In the final stage of language processing, a clinical BERT model was used to extract the sentiment in the note eg, identify if the note asserts SAR recommendation or negates SAR recommendation. This information was converted to structured data and combined with other structured data from the patient for further analysis and interpretation.

### NLP-based automatic referral extractor

All analyses were completed on the extracted data using Python (version 3.9). To identify whether a patient was recommended for SAR, we mined the text in each clinical note using the regular expression (re) library. We first standardized the text by replacing all characters that were not letters with a space. Then, we searched each clinical note with a length of at least 5 characters for the keywords “SAR” or “subacute rehabilitation.” We also searched for an indication that SAR was recommended by searching each note for “rec” (recommended) or “anticip” (anticipated). If a match was found for both the SAR and recommendation keywords, we calculated the minimum distance between their starting indices in the string. To account for negations (eg, SAR was not recommended), we implemented the Clinical Assertion/Negation Classification BERT model with the use of the transformers library^22^. This model takes text as input and classifies whether medical conditions are present, absent, or possible. Specifically, we passed the text in the note between the starting index for the first of the recommendation or SAR keyword through 5 characters past the starting index of the other keyword. We marked each clinical note as a recommendation for SAR if i) a match in the note was found for both the recommendation and SAR key terms, ii) the minimum distance between those terms was less than or equal to 40 characters, and iii) the Clinical Assertion/Negation Classification BERT model indicated an assertion, or recommendation. To identify patients that were eventually discharged to SAR, we searched the Discharge Disposition note for each patient and labeled those with the following note as transferred to SAR: “Discharged/transferred To Skilled Nursing Facility (snf) With Medicare Certification.”

### Statistical analysis

Our aim was to determine if there was a difference in outcomes for Hospital at Home patients and the control group, patients who did not participate in the Hospital at Home program. Towards this goal, we considered recommendations for SAR as well as discharges to SAR. Specifically, we summed the total number of patients who were recommended for SAR for the Hospital at Home patient group and control group. We also summed the number of patients that were recommended for SAR that were eventually discharged to SAR. For the recommendation comparison, we tested whether the proportion of patients recommended for SAR was different between the two groups using a two-tailed two proportion Z-test. We calculated the test statistic,

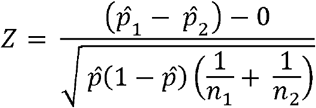

where 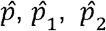 is the proportion recommended for SAR across both groups, the proportion recommended for SAR for the Hospital at Home group, and the proportion recommended for SAR for the control group, respectively, and *n*_1_ and *n* _2_ are the total number of patients in each group, respectively. We did not run this statistical test for the discharge comparison because this test requires that there are at least 10 patients who are discharged to SAR (and not discharged to SAR) for each group. Our results indicate that this requirement was not met.

## Results

We aimed to determine if outcomes for patients in Virtua’s Hospital at Home program were different from patients who did not participate in the program and remained in the hospital. Towards this goal, we extracted EMRs from patients who participated in the Hospital at Home program in 2022 (N = 272; 114 male, 158 female; age (years), m: 77.82, std: 9.95). We found that there were 43 observed unique diagnoses within this group, based on the MS_DRG codes in the records. In order to compare the Hospital at Home group to a comparable population, we limited the control group to inpatients at a Virtua hospital in 2022 who had a diagnosis (MS_DRG) that matched one of the unique diagnoses in the Hospital at Home group (N = 13879; 6366 male, 7513 female; age (years), m: 68.16, std: 20.29).

We used recommendation for SAR and discharge to SAR as metrics for outcomes following discharge from inpatient care. These metrics were chosen because they can be tracked within the Virtua Health system (for example, it would be more difficult to track re-admissions if a patient was re-admitted to a hospital outside the Virtua Health system). For each clinical note and for each patient, we used text mining and Clinical Assertion/Negation Classification BERT model^22^ (see **Methods**). After labeling each note as a recommendation to SAR (or not), we summed the number of patients for which there was at least one mention of a SAR recommendation. For the Hospital at Home group, only 40 of the 272 patients (14.7%) were recommended for SAR (**Table 1**). In contrast, 5012 of 13879 patients (36.1%) in the control group were recommended for SAR. This difference in the proportion of patients recommended for SAR across the two groups was significantly different (**Figure 2**; p < 0.001, Z = -7.30, two-tailed two proportion Z-test), with relatively far fewer Hospital at Home patients recommended for SAR.

**Table 1.** Number of patients in the Hospital at Home and control groups recommended for SAR

|  | Hospital at Home | Control |
| --- | --- | --- |
| SAR recommendation | 40 | 5012 |
| No SAR recommendation | 232 | 8867 |

**Figure 2.**
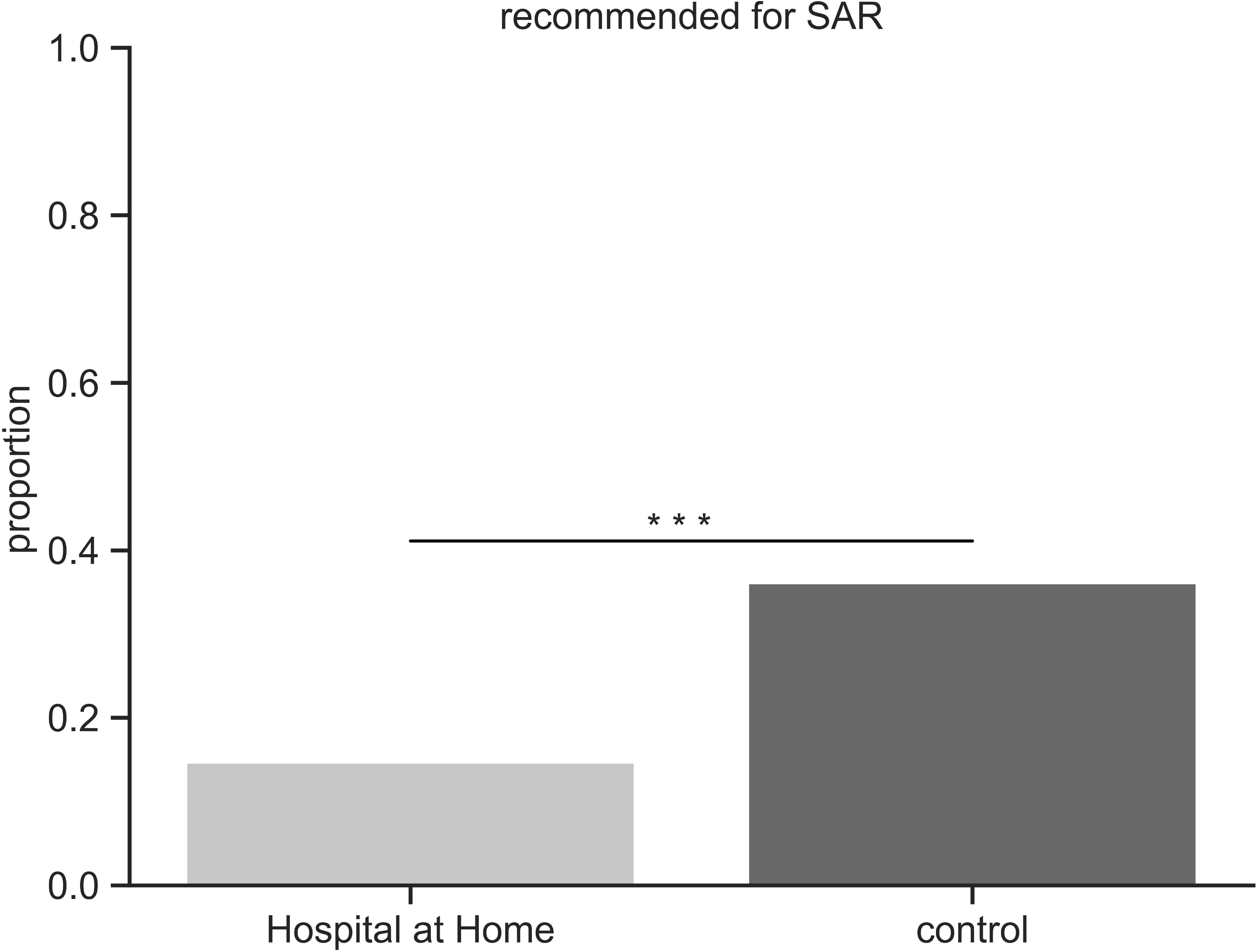
Proportion of patients in the Hospital at Home program that were recommended for SAR (0.147; N = 40 of 272) was significantly different from that in the control group (0.361; N = 5012 of 13879) (p < 0.001, Z = -7.30, two-tailed two proportion Z-test).

We were interested in knowing whether there was a significant difference in the final disposition of patients who had a recommendation for SAR between the Hospital at Home and control groups. To address this question, we searched the Discharge Disposition note for indication that the patient was transferred to SAR. Of the 40 patients in the Hospital at Home group who had a recommendation for SAR in their EMR, only 1 (proportion = 0.025) was eventually discharged to SAR (**Table 2**). In contrast, 2499 of the 5012 patients (proportion = 0.499) recommended for SAR were eventually discharged to SAR (**Figure 3**). Because only 1 patient in the Hospital at Home group was discharged to SAR, we did not perform a statistical test (N too low). However, the striking difference in the proportions of patients transferred to SAR between the groups suggests that Hospital at Home patients fared better than their in-hospital counterparts.

**Table 2.** Number of patients in the Hospital at Home and control groups discharged to SAR

|  | Hospital at Home | Control |
| --- | --- | --- |
| SAR discharge following SAR recommendation | 1 | 2499 |
| No SAR discharge following SAR recommendation | 39 | 2513 |

**Abbreviations:** CMS, Centers for Medicare & Medicaid Services; DRG, diagnosis related group; NLP, natural language processing; SAR, subacute rehabilitation.

**Figure 3.**
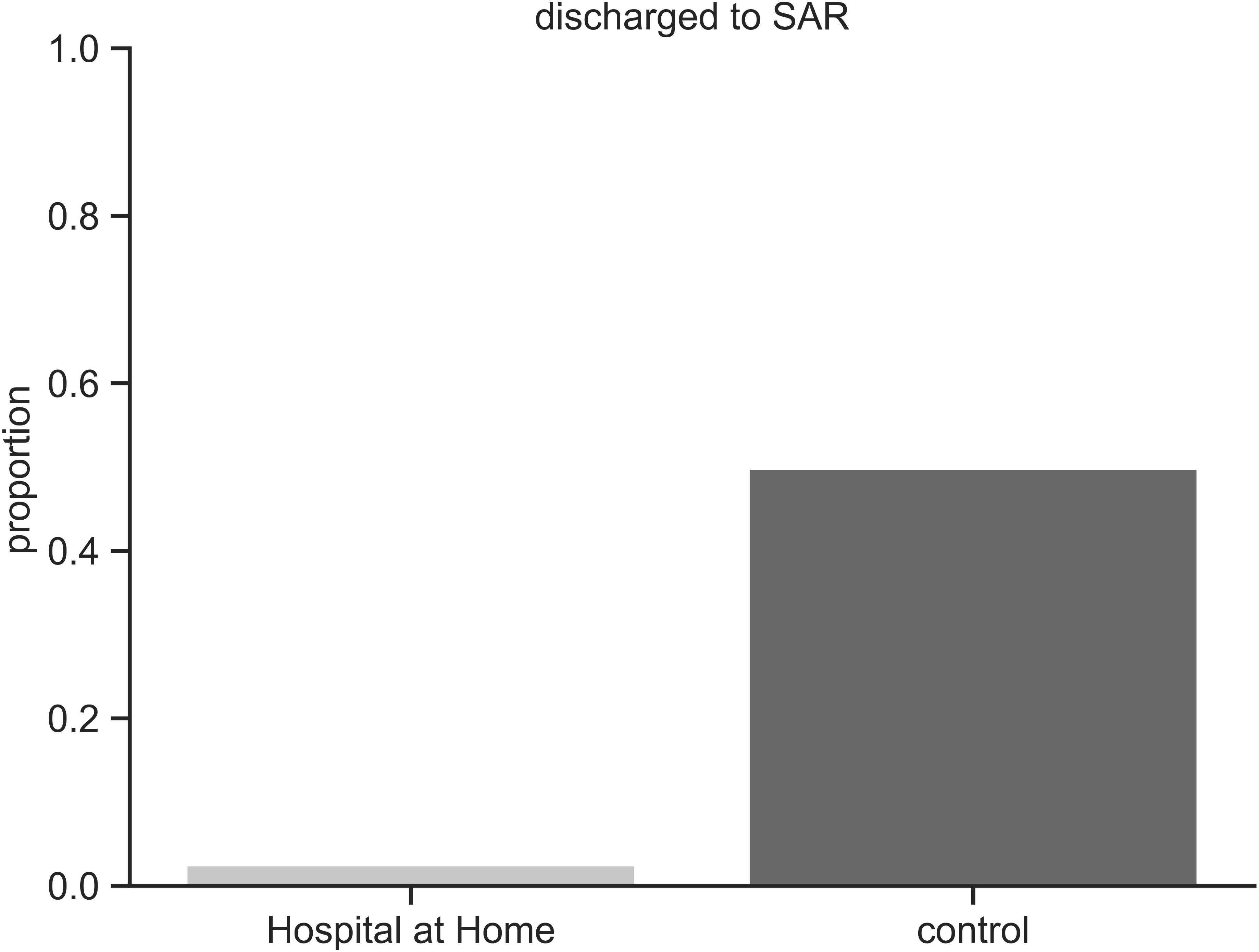
The proportion of patients in the Hospital at Home program that were both recommended and discharged for SAR (0.026; N = 1 of 40) was lower than that in the control group (0.499; N = 2499 of 5012). Because only one patient was discharged to SAR from the Hospital at Home group, this comparison did not meet the requirements for the two proportions Z-test.

## Discussion

Virtua’s Hospital at Home program admits patients from a variety of DRGs, with the most common diagnoses being pneumonia, congestive heart failure (CHF), chronic obstructive pulmonary disease (COPD), asthma, cellulitis, urinary tract infection (UTI), and COVID. Currently, all patients admitted to the Hospital at Home program are Medicare patients. Prior to being transferred home, all patients have a baseline physical therapy evaluation. Recommendations following this evaluation range from no needs to home with assistive devices, home physical therapy, or transfer to a rehabilitation facility. Of the Hospital at Home patients included in this study, 14.7% had a baseline recommendation for SAR on discharge and only 1 patient was eventually discharged to SAR. In contrast, 36.1% of patients treated in the brick-and-mortar hospital setting had a baseline recommendation for SAR, and 49% of those patients with a recommendation went to SAR on discharge.

One possible reason for the lower rate of patients going to SAR in the Hospital at Home group compared to the brick-and-mortar inpatient group may be due to differences in mobility. We notice that patients in the Hospital at Home program benefit from increased and early mobility, as assessed from remote monitoring kits tracking step counts. While at home, patients can liberalize their ambulation. They are not restricted to the hospital setting, which in most cases means staying in a semi-private room. Being at home also correlates with decreased hospital-acquired delirium as the patients are familiar with the layout and nuances of their home. Furthermore, at home, patients can be with their loved ones and pets, and sleep in their own beds. In addition to these liberties, patients likely feel more secure knowing that we are monitoring them 24/7 with remote monitoring kits, providing us with valuable information.

We believe that patients in the Hospital at Home program experience early mobility, better sleep cycles and decreased delirium, all of which translate to less deconditioning and subsequently no need for SAR on discharge.

Currently we are limited to admitting only Medicare patients to our program. We have no reason to believe that these outcomes are specific to Medicare patients and predict that these outcomes would hold if expanded to a broader patient population.

## Conclusion

We observed that patients in Virtua’s Hospital at Home program were recommended for SAR (14.7%) at less than half of the rate of inpatients in the traditional brick-and-mortar hospital (36.1%). This reduced need for SAR for the Hospital at Home patients is likely due to relatively increased mobility, better sleep, and decreased rate of delirium in the home compared to the brick-and-mortar hospital. The results from this study suggest that the Hospital at Home program is an attractive alternative to traditional inpatient care for patients who meet certain clinical criteria.

## Data Availability

Data produced in the present study are not available in order to comply with agreements made with the ethics committee (IRB).

